# Burden of Fecal Colonization with Extended-Spectrum Beta-Lactamase Producing *Enterobacteriaceae* among Food handlers in Africa: A Systematic review and meta-analysis

**DOI:** 10.64898/2025.12.30.25343138

**Authors:** Amanuale Zayede, Yitayih Wondimeneh, Eshet Gebrie, Mitkie Tigabie, Henok Worku, Biruktawit Abebe, Haymanot Til, Sirak Biset, Mucheye Gizachew

## Abstract

**Background:** Antimicrobial resistance particularly from extended-spectrum β-lactamase-producing *Enterobacteriaceae* (ESBL-PE) is a critical global public health threat, contributing significantly to infection-related mortality. The human intestinal tract is a key reservoir for ESBL-PE, and exposure to antibiotics can disrupt the gut microbiota, thereby making it easier for these pathogens to colonize the intestine. Colonized individuals are at risk of developing subsequent infections and can silently transmit these resistant bacteria to others. This transmission risk is particularly significant in food handlers, who can inadvertently spread pathogens to consumers, posing a substantial food safety hazard. In Africa, the rapid spread of ESBL-PE is exacerbated by a lack of regular surveillance and antibiotic stewardship programs. While primary studies on ESBL-PE exist, a comprehensive synthesis of data specific to the food handler population is lacking. Therefore, this systematic review and meta-analysis aimed to determine the pooled fecal colonization rate of ESBL-PE among food handlers in Africa to inform public health interventions.

**Materials and Methods:** This review was conducted according to a protocol registered in PROSPERO (ID: CRD420251075141). A systematic search was performed from September 5 to 15, 2025 in PubMed, Google Scholar, and Hinari/Research4Life to identify relevant studies. The methodological quality of the included studies was appraised using the Joanna Briggs Institute (JBI) critical appraisal tool. Data were extracted into Microsoft Excel 2019 and analyzed with Stata software version 17. Given significant heterogeneity among the studies (I² = 98.3%, *p* < 0.001), a random-effects meta-analysis model (DerSimonian and Laird) was used to calculate the pooled prevalence. To investigate the substantial heterogeneity, we performed subgroup analysis. Furthermore, sensitivity analysis was conducted to assess the influence of individual studies on the overall results. The potential for publication bias was assessed visually with a funnel plot and statistically using Egger’s test. Finally, results were presented via text, figures, and tables.

**Results:** The meta-analysis incorporated nine studies with publication year ranged between 2012 and 2023, involving 4,061 participants. The pooled fecal colonization rate of ESBL-PE among food handlers in Africa was 23.64% (95% CI: 15.3, 31.94%). A high degree of heterogeneity was observed (I² = 98.3%, *p* < 0.001). The most prevalent ESBL-PE species identified was *E. coli*, with a pooled prevalence of 85.83% (95% CI: 79.97, 91.69%, I^2^ = 95.56%, *p* <0.001), followed by *Klebsiella* species at 23.92% (95% CI: 18.48, 29.35%, I^2^ = 0.00%, *p* = 0.65)

**Conclusion and recommendations:** This meta-analysis establishes that approximately one in four food handlers in Africa are colonized with ESBL-PE, indicating a substantial reservoir for community transmission. This colonization risk facilitates the spread of antimicrobial resistance and can lead to severe, hard-to-treat infections. We therefore recommend implementing targeted public health measures, including routine screening, antimicrobial stewardship, and strict infection control protocols for food handlers to mitigate this threat.

## Introduction

*Enterobacteriaceae* are a major family of Gram-negative bacteria that include notable pathogens such as *Escherichia*, *Klebsiella*, *Salmonella*, *Shigella*, and *Enterobacter* species. These organisms are a common cause of diverse infections in both healthcare and community settings [1, 2]. Of particular concern are extended spectrum beta lactamase-producing *Enterobacteriaceae* (ESBL-PE), which the World Health Organization (WHO) has classified them as critical-priority pathogens due to their high resistance to third generation cephalosporins [3]. *Escherichia coli (E. coli) and Klebsiella pneumoniae (K. pneumoniae)* are two common species of *Enterobacteriaceae* that possess pathogenic capabilities and carry ESBL-encoding genes [4]. The Infectious Diseases Society of America (IDSA) has identified them as two of the six pathogens for which new drugs are urgently required to address the issue of resistance development [5]. According to the Global antimicrobial resistance (AMR) and Use Surveillance System (GLASS) Report 2022, resistance rates to meropenem and third-generation cephalosporins in *E. coli* infections increased by more than 15% in 2020 compared to 2017 [6].

Experts predict that by 2050, AMR could cause approximately 1.91 million deaths annually worldwide. Without effective interventions, cumulative AMR-related deaths may reach 39.1 million between 2025 and 2050, demanding urgent global action to mitigate this crisis [7]. A growing public health concern is the widespread intestinal carriage of ESBL-PE, which contributes to infections in both community and healthcare environments. Even when asymptomatic, colonization with these resistant pathogens remains a serious issue, heightening the risk of difficult to treat infections [8].

The production of beta-lactamases, enzymes that hydrolyze and inactivate beta-lactam antibiotics, is one of the major mechanism of AMR in bacteria. The ESBL are a particularly dangerous subset of these enzymes that can break down third-generation cephalosporins (e.g., cefotaxime, ceftriaxone, ceftazidime) and monobactams (e.g., aztreonam) [4, 9]. This resistance is acquired by plasmid-mediated mutation encoding for the parent enzymes, either by amino acid substitution in the active site such as the case for Temonieara (TEM) or sulfhydryl reagent variable (SHV; class A) enzymes or by inter bacteria gene transfer like the case of cephalosporinases (class C enzymes) [10, 11]. The ESBL-PE are among the most urgent AMR threats, as identified by the United states (US) Centers for Disease Control and Prevention (CDC) [12]. These pathogens severely limit treatment options, as they often require the last resort antibiotics such as carbapenems [13]. The spread of these resistant bacteria leads to higher mortality rates, prolonged hospital stays, and increased healthcare costs [14, 15].

The transmission of ESBL-PE occurs through multiple reservoirs, including colonized patients, contaminated medical equipment and food products [16]. Additionally, the gut microbiota can serve as a reservoir for resistance genes, facilitating their transfer to pathogenic strains [17].

A global meta-analysis investigating fecal colonization with ESBL-PE among hospitalized patients, which included data from Africa, reported a pooled prevalence of 45.6% [18]. Further evidence from Africa indicates substantial colonization rates in both community settings (18%) and among patients at hospital admission (32%) [19]. Despite this established burden, there remains a significant lack of synthesized data specific to food handlers, a key demographic for public health surveillance. Therefore, this systematic review and meta-analysis was conducted to determine the pooled fecal colonization rate of ESBL-PE among food handlers across Africa.

## Materials and Methods

### Study protocol

This systematic review and meta-analysis followed the Preferred Reporting Items for Systematic Reviews and Meta-Analyses (PRISMA) 2020 checklist and guidelines (**S1 Table**) [20]. We began by registering our study protocol with PROSPERO (Registration ID: CRD420251075141).

### Search strategy

A systematic literature search was performed from September 5 to 15, 2025, in PubMed, Google Scholar and Hinari/ Research4life. To maximize the scope of the review, we also manually examined the reference lists of all identified relevant studies to locate any additional publications, gray literatures, and unpublished studies from preprint servers that were not captured by the initial database search. The search strategy was built using a combination of appropriate Medical Subject Headings (MeSH) terms and keywords related to the core concepts of our research question. These terms were strategically combined using Boolean operators: "OR", and "AND" to focus the search.

An example of the full search string used for PubMed is provided for transparency: (((((((((("Fecal carriage") OR ("Fecal colonization")) OR ("Gut carriage")) OR ("Gut colonization")) OR ("Intestinal colonization")) OR ("Intestinal carriage")) OR (Colonization)) OR (Carriage))) AND (((((("Extended spectrum beta lactamase") OR ("Extended spectrum β lactamase")) OR (ESBL)) OR (ESβL)) OR (Beta-Lactamase)))) AND (((((((((((((((Enterobacteriaceae) OR (Enterobacterales)) OR ("Gram-Negative Bacteria")) OR ("Gram-Negative Bacteria"[Mesh])) OR (Enterobacteriaceae[Mesh])) OR (Klebsiella)) OR (Klebsiella[Mesh])) OR ("Escherichia coli")) OR ("Escherichia coli"[Mesh])) OR (E.coli)) OR (Shigella)) OR (Shigella[Mesh])) OR (Salmonella)) OR (Salmonella[Mesh])) AND (((((((Africa) OR (Africa[Mesh])) OR ("Sub Saharan Africa")) OR ("Sub Saharan Africa")) OR ("Africa South of the Sahara"[Mesh]))) AND ((((((Africa) OR (Africa[Mesh])) OR ("Sub Saharan Africa")) OR ("Sub Saharan Africa")) OR ("Africa South of the Sahara"[Mesh]))))) **(S2 Table)**

### Eligibility criteria

We applied the CoCoPop (Condition, Context, and Population) approach to determine the inclusion and exclusion criteria. In this framework, the prevalence of ESBL-PE was defined as the condition (Co), food handlers providing fecal specimens or rectal swabs represented the population (Pop), and Africa was designated as the context (Co).

#### Inclusion criteria

We included observational studies (cross-sectional, case-control, and cohort designs) that investigated the prevalence or factors associated with laboratory confirmed ESBL-PE carriage. The population of interest was adult food handlers (≥18 years) employed in any food service setting, including restaurants, street vending, and institutional catering services across any African country. Eligible studies had to confirm ESBL-PE status through laboratory analysis of stool or rectal swab samples. There were no restrictions on the publication date. To reduce publication bias, gray literature and unpublished studies from preprint servers and conference abstracts were also considered.

#### Exclusion criteria

Reviews, case reports, case series, animal studies, and studies not based on stool samples were excluded. Research focusing on food handlers with acute diarrheal illness at sampling was also excluded to avoid capturing transient colonization.

### Study selection

All records identified through the database searches were imported into Endnote X9 for reference management, and duplicate were removed. The study selection process was carried out in two phases using the predefined CoCoPop framework. First, two reviewers (AZ and MG) independently screened the titles and abstracts of all identified citations against the eligibility criteria. Second, the full texts of the potentially relevant studies were retrieved and assessed independently by the same reviewers for final inclusion. Any disagreements between the reviewers at either stage were resolved through discussion or by consulting a third and fourth reviewers (YW and SB).

### Quality appraisal

The methodological quality of the included studies was assessed using the Joanna Briggs Institute (JBI) critical appraisal checklists [21]. Two reviewers (AZ and MG) independently applied the relevant JBI checklist to each study. These checklists contain nine items, and studies were scored as follows: 0-4 (low quality), 5-7 (moderate quality), and 8-9 (high quality). Only studies achieving a score of 5 or higher (i.e., moderate or high quality) were incorporated into the final analysis. Any discrepancies between the reviewers’ assessments were resolved through discussion or by consulting a third and fourth reviewers (YW and SB).

### Outcome variable measurement

The primary outcome for this meta-analysis was the pooled prevalence of fecal colonization with ESBL-PE among adult food handlers across Africa. For each included study, the prevalence was calculated as the proportion of tested individuals with a laboratory-confirmed positive result. Specifically, the number of ESBL-PE positive participants was divided by the total number of participants screened in that study, and the result was multiplied by 100 to yield a percentage.

### Data extraction and management

All records identified from the electronic databases were combined and properly exported to Endnote version 9.2 for management. After merging the records into a single library, duplicate articles were systematically identified and removed. The remaining unique records were then screened by two independent reviewers (AZ and MG), using the predefined inclusion and exclusion criteria.

Subsequently, a standardized data extraction form was developed in Microsoft Excel 2019, following the PRISMA guidelines. Two authors (AZ and MG) independently extracted the following data from each included full-text article: primary author, publication year, study design, study period and setting, geographical region, sample size, number of cases, ESBL-PE detection method, and the rates of overall ESBL-PE colonization as well as individual bacterial strains. Any discrepancies in the extracted data were resolved through discussion until a consensus was reached.

### Data synthesis and statistical analysis

All statistical analysis were performed using Stata software version 17. The extracted data were summarized using text and tables. To calculate the pooled prevalence, we generated forest plots, which provide a visual summary of each study’s effect size and confidence interval (CI). The degree of statistical heterogeneity among the included studies was quantified using the I² statistic. The I² values were interpreted as follows: less than 25% represented low heterogeneity, 25% to 50% indicated moderate heterogeneity, 50% to 75% represented substantial heterogeneity, and values exceeding 75% signified considerable heterogeneity. The statistical significance of the observed heterogeneity was determined by the *p* value of the Cochrane Q statistic, and *p*-value of less than 0.05 was evidence of heterogeneity [22]. Given the anticipated heterogeneity, a DerSimonian and Laird random-effects model was employed to estimate the overall prevalence of ESBL-PE colonization in Africa. The final pooled estimates for ESBL-PE prevalence are presented with 95% CI in forest plots and tables.

### Subgroup and sensitivity analysis

Given the significant statistical heterogeneity identified among the included studies, we conducted in-depth analysis to explore its potential sources and assess the robustness of our findings. A subgroup analysis was performed by stratifying the studies based on three pre-specified factors: the geographic region in Africa (Eastern, Western, and Northern,), study year (2012-2019, and 2020-2025), and the specific laboratory method used for ESBL-PE detection double disk synergy test (DDST), and combination disk test (CDT). Additionally, a leave-one-out sensitivity analysis was conducted to assess the influence of individual studies on the overall pooled prevalence of ESBL-PE.

### Publication bias

The potential for publication bias and small study effects across the included studies was assessed using both graphical and statistical methods. A funnel plot was generated and visually inspected for symmetry; a symmetrical, inverted funnel shape suggested a low likelihood of publication bias. To complement this visual assessment, Egger’s regression test was employed. A *p*-value greater than 0.05 from Egger’s test was interpreted as indicating no statistically significant publication bias [23].

## Results

### Selection of studies

The systematic search across scientific databases initially yielded 752 records. Following the removal of 37 duplicates, 715 unique articles remained for screening. After a review of the titles and abstracts, 672 records were excluded as irrelevant. The full texts of the remaining 43 articles were then assessed for eligibility based on the predefined inclusion criteria and quality appraisal. Finally, 9 studies were eligible and included in the final meta-analysis, as detailed in the PRISMA flow diagram (**Fig 1**).

**Fig 1:**
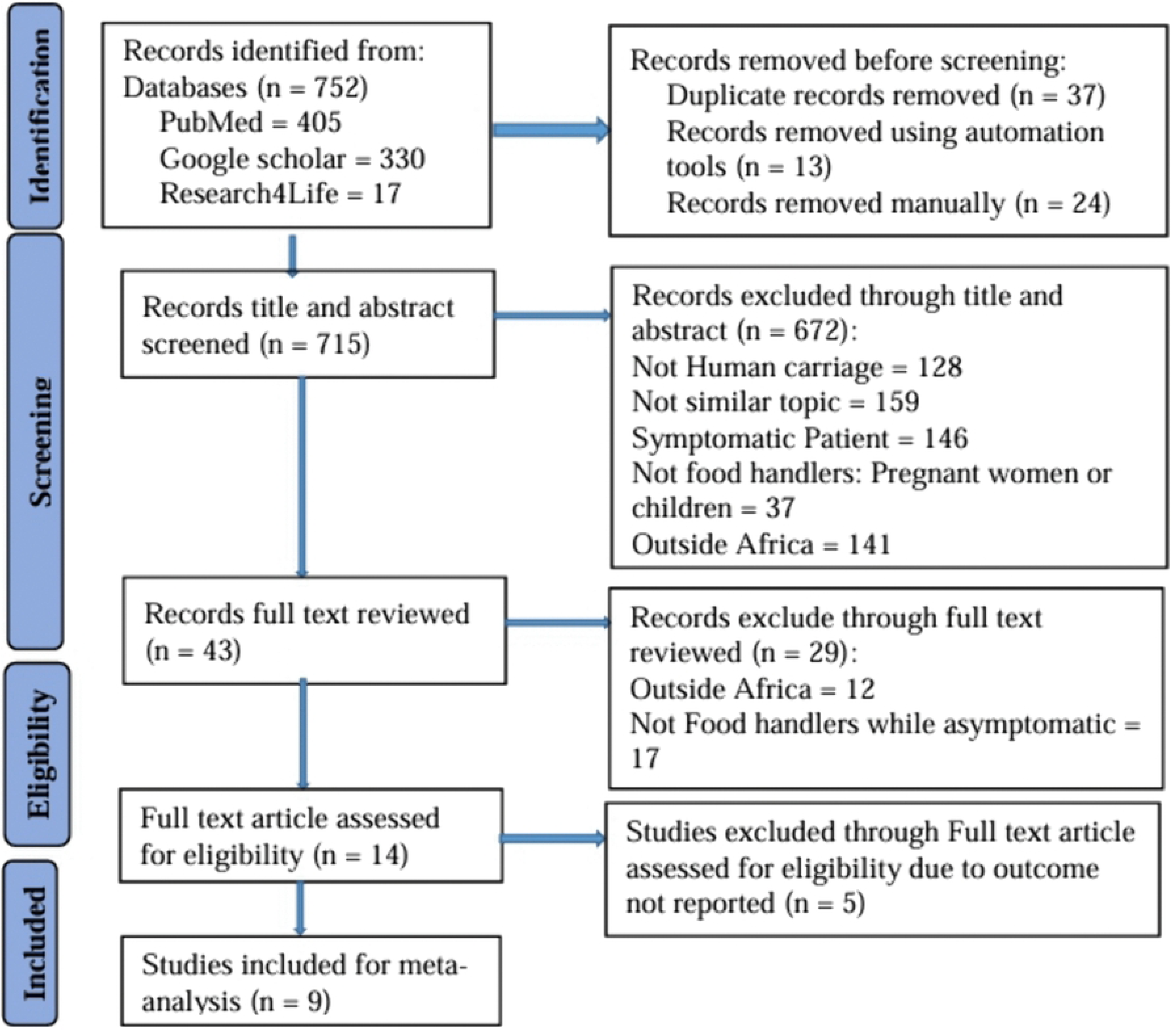
PRISMA flow diagram of the study identification and selection process for the systematic review and meta-analysis of ESBL-PE colonization rate among food handlers in Africa, 2025

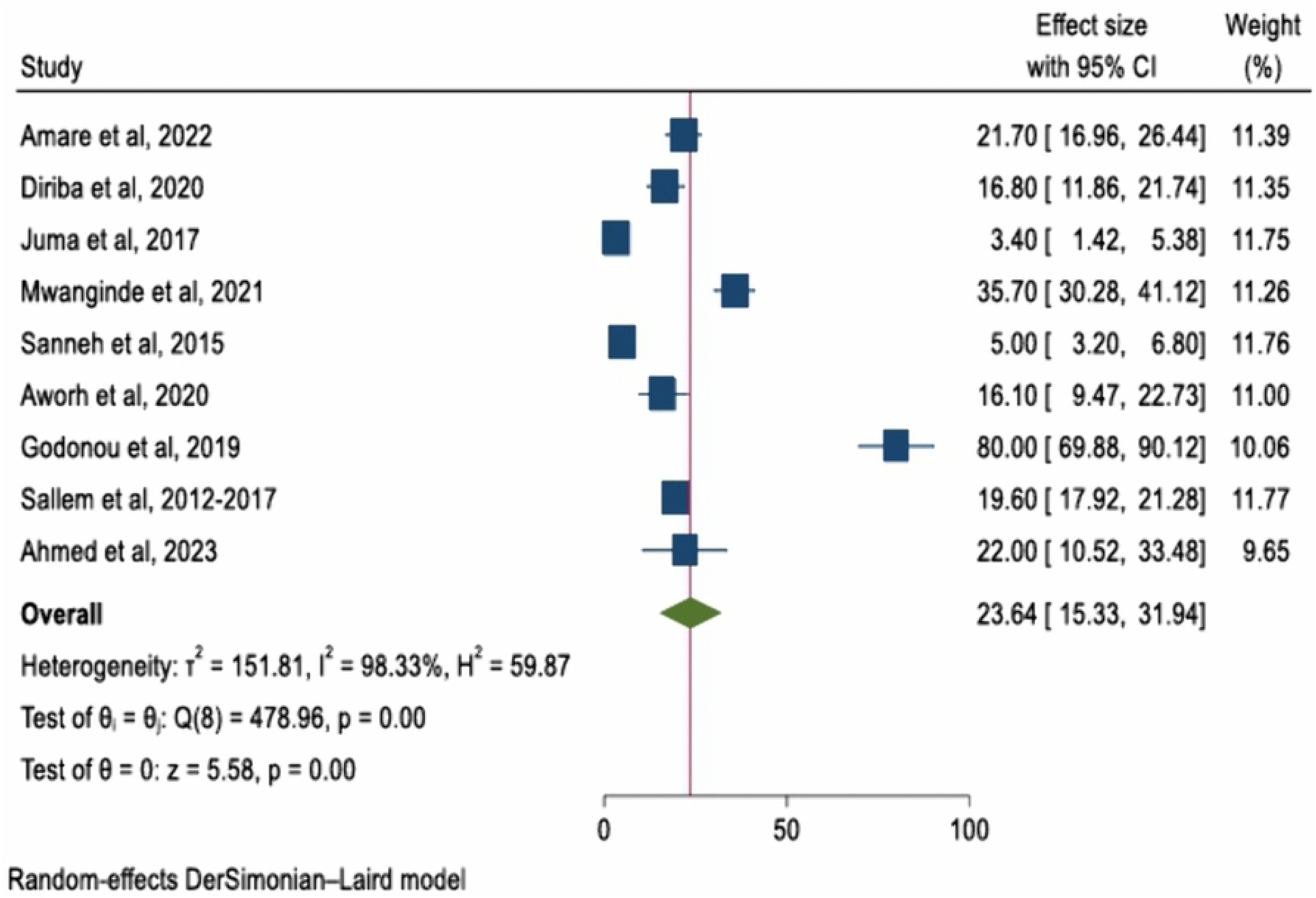

### Summary of quality appraisal of included studies

Regarding the studies that met the quality criteria, 5 out of 9 (55.6%) had a good quality, while 4 (44.4%) had a moderate quality. No studies were excluded due to a low quality (**Table 1**).

**Table 1:** Quality appraisal of included studies evaluating the fecal colonization rate of ESBL-PE among Food handlers in Africa, 2025.

| Author, Year | Study design | Q1 | Q2 | Q3 | Q4 | Q5 | Q6 | Q7 | Q8 | Q9 | Score out of 9 | Remark |
| --- | --- | --- | --- | --- | --- | --- | --- | --- | --- | --- | --- | --- |
| Ahmed et al, 2023 [38] | CS | UN | No | No | Yes | Yes | Yes | Yes | Yes | Yes | 6 | Included |
| Amare et al, 2022 [39] | CS | Yes | Yes | Yes | Yes | Yes | Yes | Yes | Yes | Yes | 9 | Included |
| Aworh et al, 2020 [40] | CS | No | Yes | No | Yes | Yes | Yes | Yes | Yes | Yes | 7 | Included |
| Diriba et al, 2020 [41] | CS | Yes | Yes | UN | No | Yes | Yes | Yes | Yes | Yes | 7 | Included |
| Godonou et al, 2019 [26] | CS | UN | Yes | No | Yes | Yes | Yes | Yes | Yes | Yes | 7 | Included |
| Juma et al, 2017 [24] | CS | Yes | Yes | Yes | Yes | Yes | Yes | Yes | Yes | Yes | 9 | Included |
| Mwanginde et al, 2021 [42] | CS | Yes | Yes | Yes | Yes | Yes | Yes | Yes | Yes | Yes | 9 | Included |
| Sallem et al, 2012-2017 [43] | CS | Yes | No | Yes | Yes | Yes | Yes | Yes | Yes | Yes | 8 | Included |
| Sanneh et al, 2015 [25] | CS | Yes | Yes | Yes | Yes | Yes | Yes | Yes | Yes | Yes | 9 | Included |

**Abbreviations:** CS=Cross-sectional; UN=unclear; Q=Question.
**Note:** The overall score is calculated by counting the number of Yes's in each row.
The following quality appraisal questions were for cross-sectional studies: Q1: Was the sample frame appropriate to address the target population? Q2: Were study participants sampled in an appropriate way? Q3: Was the sample size adequate? Q4: Were the study subjects and the setting described in detail? Q5: Was the data analysis conducted with sufficient coverage of the identified sample? Q6: Were valid methods used for the identification of the condition? Q7: Was the condition measured in a standard reliable way for all participants? Q8: Was there appropriate statistical analysis? And Q9: Was the response rate adequate, and if not, was the low response rate managed appropriately were for cross-sectional studies (**Table 1**).

### Characteristics of the studies included in the systematic review and meta-analysis

This systematic review and meta-analysis incorporated nine cross-sectional studies, with publication year ranged between 2012 and 2023, encompassing a total of 4,061 participants. The sample sizes of the individual studies varied considerably, ranging from 50 to 2,135. Regarding ESBL-PE detection methods, the majority of studies (seven) utilized the DDST, while the remaining two employed the CDT. Geographically, the studies were distributed as follows: four from Eastern Africa, three from Western Africa, and two from Northern Africa. In terms of reported outcomes, seven studies specifically documented ESBL-producing *E. coli*, while only four studies reported ESBL-producing *Klebsiella* species. Furthermore, multidrug resistance (MDR) among ESBL-PE isolates was clearly detailed in only three studies (**Table 2**).

**Table 2:**
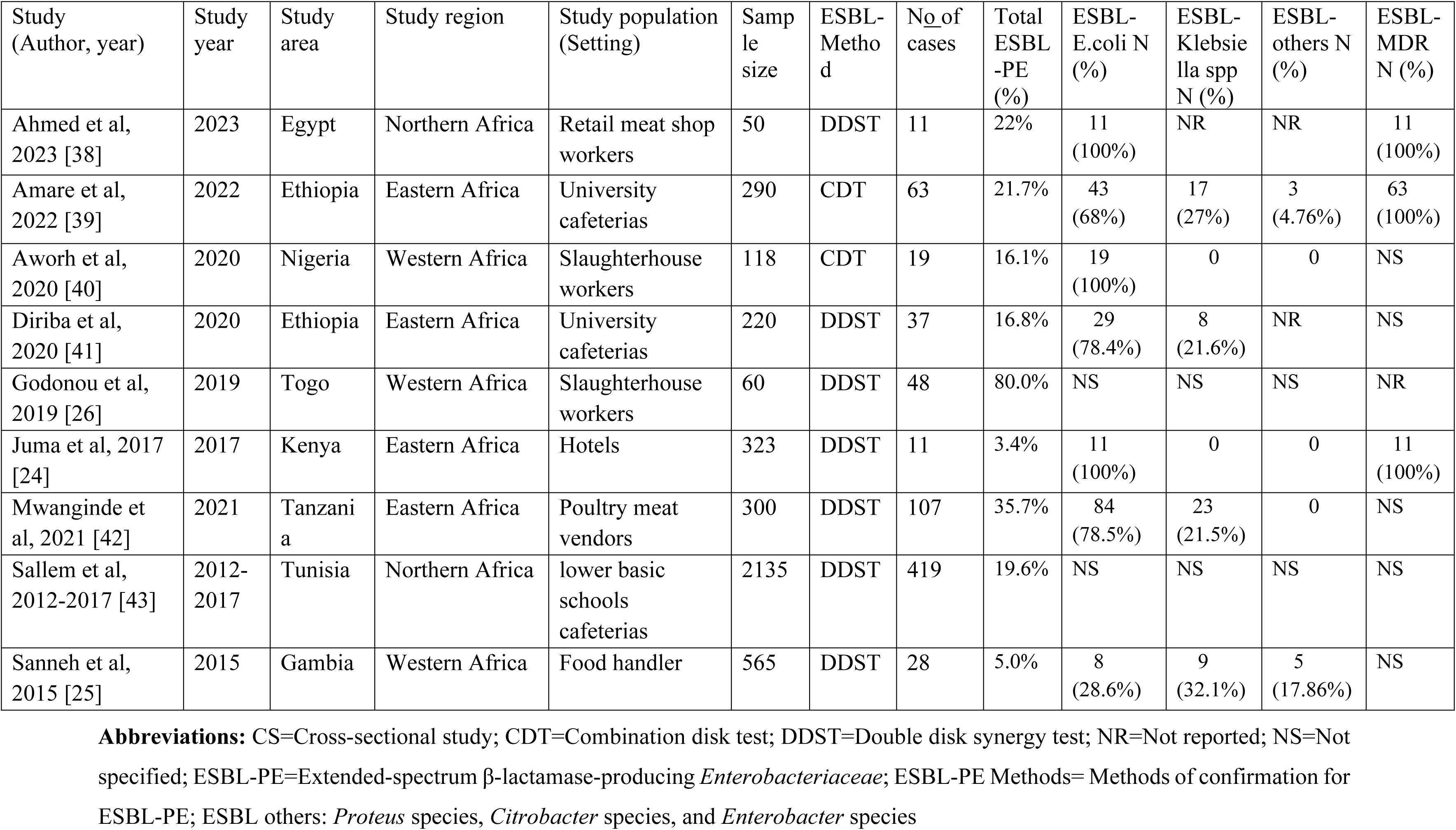
Characteristics of the individual studies included in the meta-analysis of ESBL-PE among Food handlers in Africa, 2025.

### Pooled Colonization rate of ESBL-PE among food handlers in Africa

The meta-analysis on the fecal colonization of ESBL-PE among food handlers in Africa gives a pooled prevalence of 23.64%. The nine included studies reported a prevalence ranging from 3.4% to 80.0% (**Fig 3**). Furthermore, the meta-analysis on the fecal colonization of ESBL-producing *E. coli* among food handlers in Africa gives a pooled prevalence of 85.83%. Only seven studies were reported ESBL-producing *E. coli* (**Fig 4**). Additionally, the meta-analysis on the fecal colonization of ESBL- producing *Klebsiella* species among food handlers in Africa gives a pooled prevalence of 23.92%. Only four studies were reported ESBL-producing *Klebsiella* species (**Fig 5**).

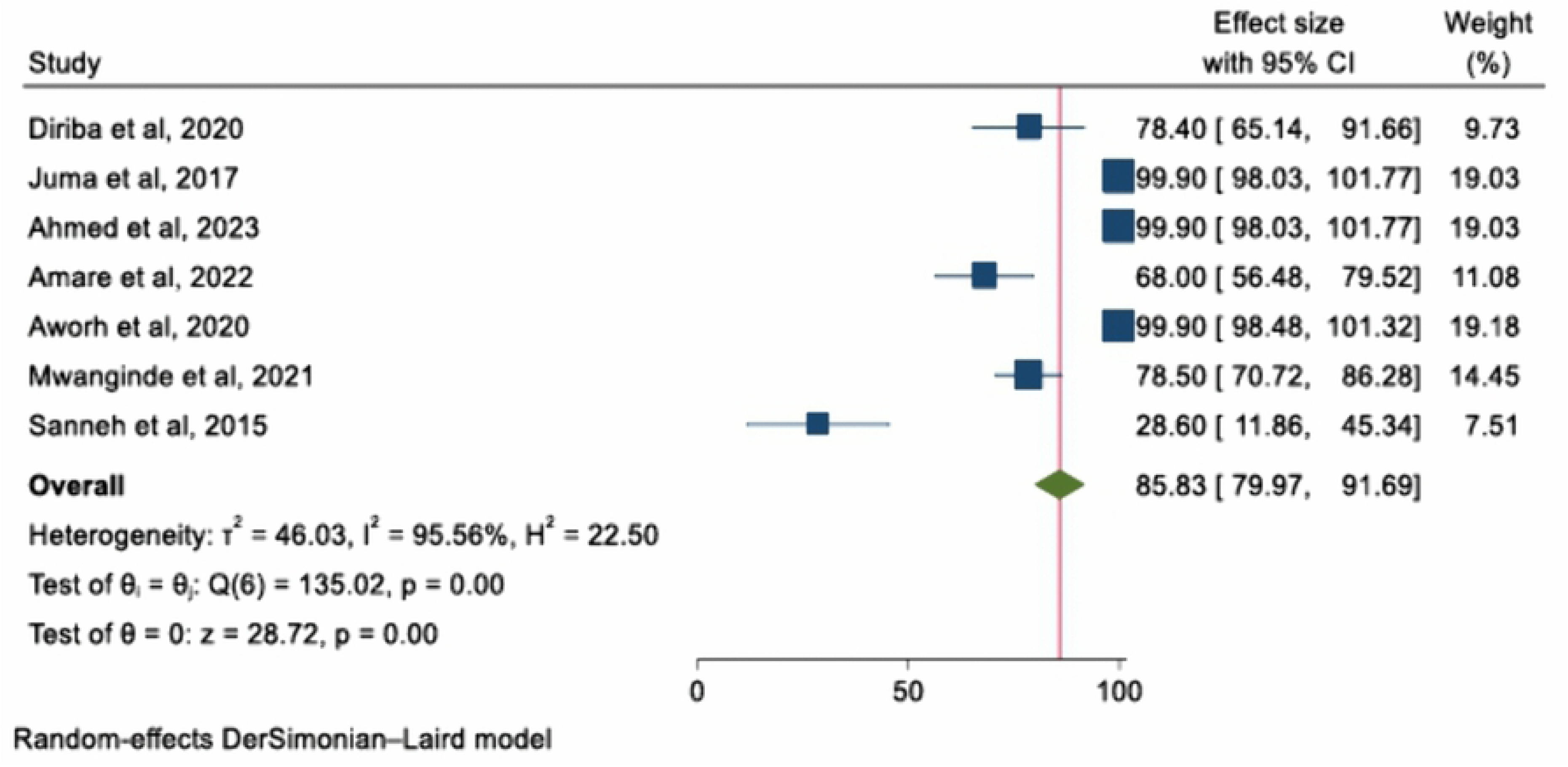

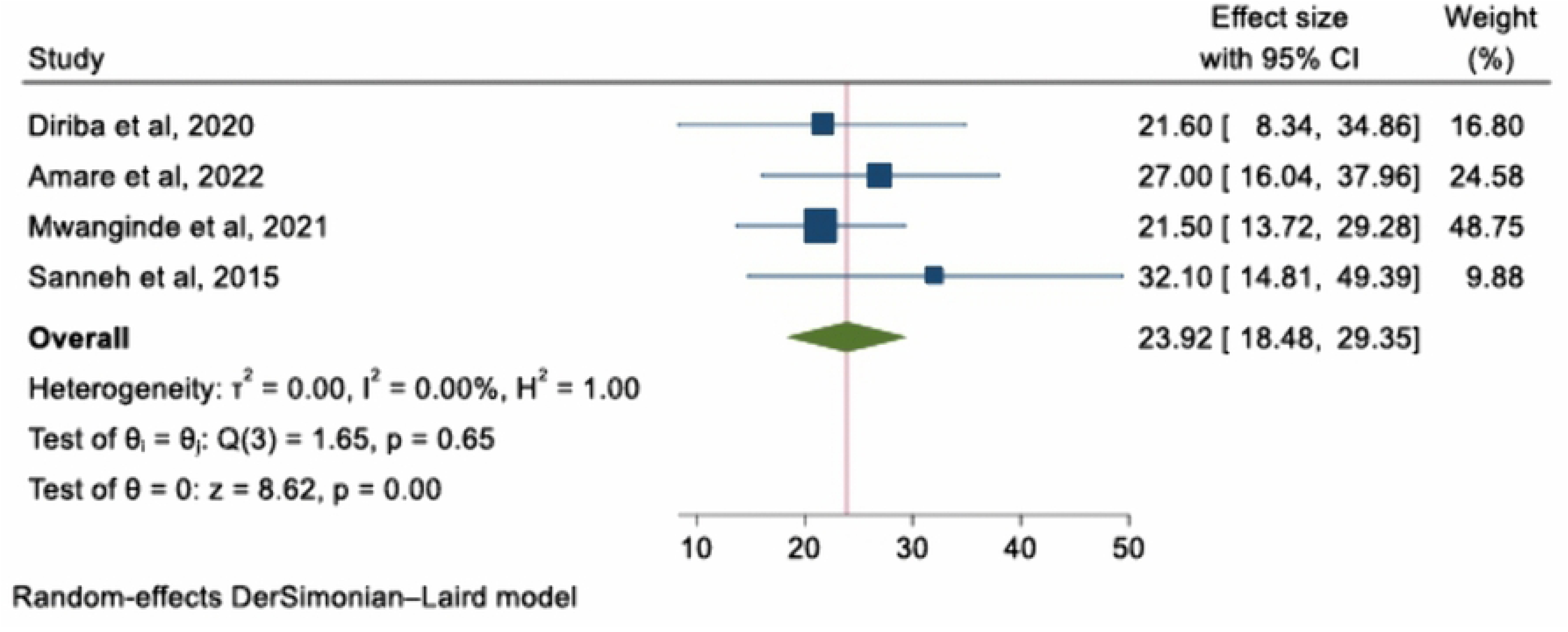

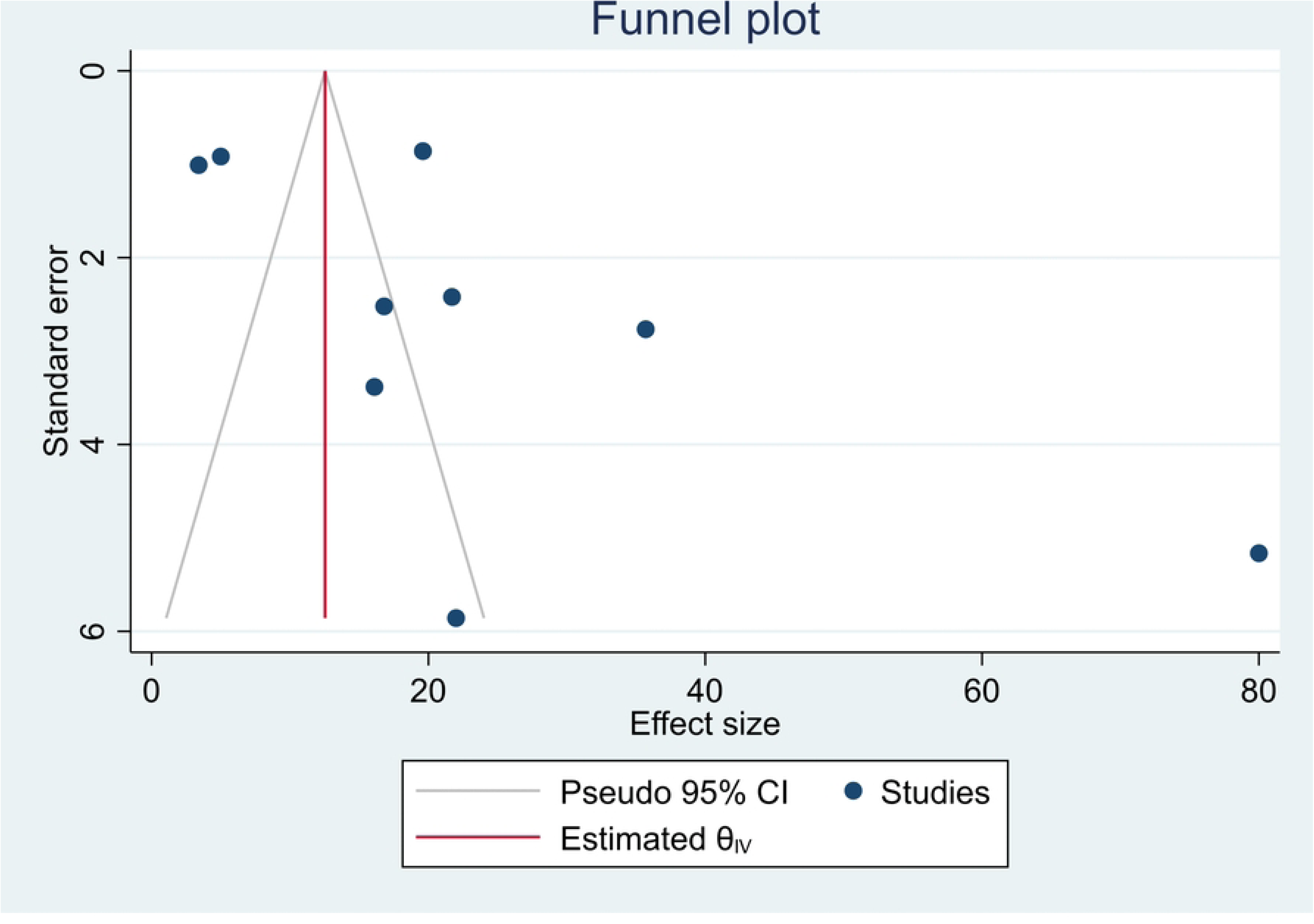

The forest plot visually represents these findings. Each study is depicted by a blue square, indicating its individual point estimate (colonization rate), and a horizontal line, representing its 95% confidence interval. The width of these lines reflects the precision of each estimate, with wider lines indicating greater uncertainty. A diamond at the bottom summarizes the overall pooled estimate, its center marks the point prevalence, while its width shows the 95% CI for the combined result **(Fig 2-4)**.

According to this meta-analysis, the pooled colonization rate of ESBL-PE among food handlers in Africa was 23.64% (95% CI: 15.3, 31.94%). A very high degree of statistical heterogeneity was observed among the study results (I² = 98.3%, *p* <0.001) (**Fig 2**), suggests that the true prevalence varies considerably across the included studies. Therefore, a random effects model was used to estimate the average effect size (prevalence). Furthermore, to explore the potential sources of this heterogeneity, subgroup and sensitivity analyses were performed **(Table 3 and 7).**

**Table 3:**
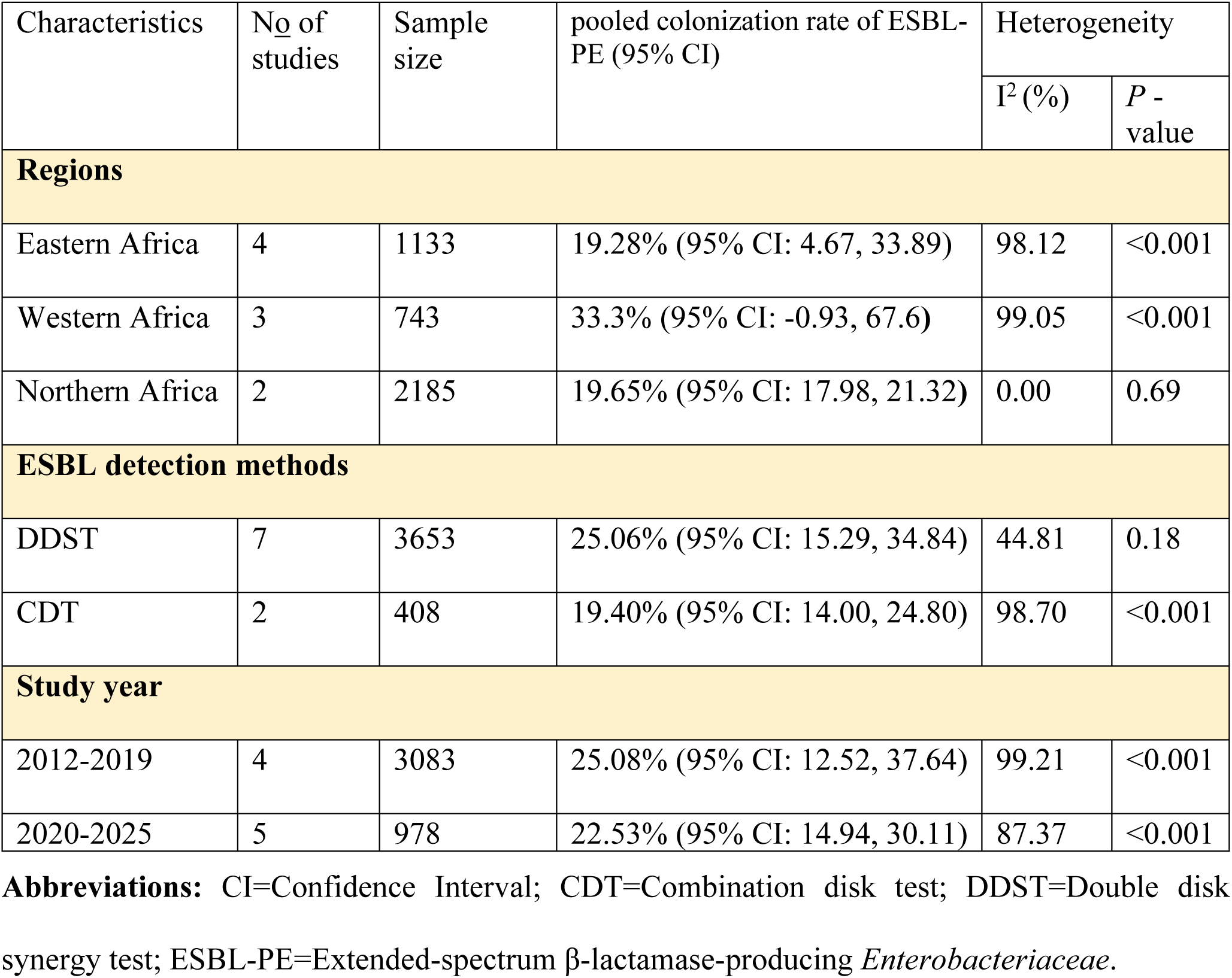
Subgroup analysis of ESBL-PE among Food handlers in Africa, 2025.

Significant variation was evident across the individual studies. The lowest colonization rates were reported in Kenya (3.4%) [24] and Gambia (5%) [25], while the highest rate was found in a study from Togo (80%) [26].

The most prevalent ESBL-PE species identified were *E. coli*, with a pooled prevalence of 85.83% (95% CI: 79.97, 91.69%, I^2^ = 95.56%, *p* <0.001) (**Fig 3**), followed by *Klebsiella* species at 23.92% (95% CI: 18.48, 29.35%, I^2^ = 0.00%, *p* = 0.65) (**Fig 4**), and ESBL-PE other than *E. coli* and *Klebsiella* species at 9.58% (95% CI: -2.80, 21.97%, I^2^ = 65.28%, *p* = 0.09) (**S1 Fig**). In terms of MDR, only three studies reported it among ESBL-PE with 100%.

### Subgroup analysis of the ESBL-PE colonization rate

In an effort to explain the substantial heterogeneity observed across the studies, we performed a subgroup analysis based on three key variables: geographic region, diagnostic method for ESBL-PE confirmation, and the study period. The analysis revealed notable regional differences. The highest pooled colonization rate of ESBL-PE was in Western Africa at 33.3% (95% CI: -0.93, 67.6), followed by Northern Africa at 19.65% (95% CI: 17.98, 21.32), and eastern Africa at 19.28% (95% CI: 4.67, 33.89). When stratified by detection method used for ESBL production, studies using the DDST reported a higher pooled prevalence of 25.06% (95% CI: 15.29, 34.84) compared to the 19.40% (95% CI: 14.00, 24.80) found in studies using the CDT. Subgroup analysis in terms of study year showed a slightly higher prevalence in studies conducted between 2012 and 2019, at 25.08% (95% CI: 12.52, 37.64), compared to those from 2020 to 2025, which had a prevalence of 22.53% (95% CI: 14.94, 30.11), as indicated in **(Table 3 and S2 Figs)**

### Publication bias

Publication bias was assessed to determine whether the selected studies represented the original population or were influenced by bias related to published and unpublished studies. The ESBL-PE analysis revealed an asymmetric funnel plot, indicating the presence of publication bias (**Fig 6**). Similarly, Egger’s test showed significant publication bias (*p* = 0.003) (**Table 4)**.

**Table 4:** Egger’s test for the pooled estimates of the colonization rates of ESBL-PE among Food handlers in Africa, 2025.

| ESBL-PE | No of studies | pooled colonization rate of ESBL-PE (95% CI) | Heterogeneity |  | Eggers regression test |  |
| --- | --- | --- | --- | --- | --- | --- |
|  |  |  | I<sup>2</sup> (%) | P - value | P - value | 95% CI |
|  | 9 | 23.64% (95% CI: 15.3, 31.94%) | 98.3% | <0.001 | 0.003 | 3.67, 12.42 |

**Abbreviations:** CI=Confidence Interval; ESBL-PE=Extended-spectrum β-lactamase-producing *Enterobacteriaceae*.

#### Trim-and-fill analysis of the pooled colonization rate of ESBL-PE

To address publication bias, a trim-and-fill analysis was performed. Without imputing data on the left, the analysis found that the pooled colonization rate of ESBL-PE among food handlers in Africa remained stable at 23.64% (95% CI: 15.3, 31.94%) **(Table 5 and S4 Figs).** Conversely, when analyzing 12 studies with three data points imputed on the right, the pooled colonization rate was slightly higher at 32.23% (95% CI: 17.31, 47.14%) (**Table 6 and S4 Figs**).

**Table 5:** Nonparametric trim-and-fill analysis of publication bias imputing on the left.

| Nonparametric trim-and-fill analysis of publication bias |  |
| --- | --- |
| Linear estimator, imputing on the left |  |
| Iteration |  |
| Parameter | Value |
| No of studies | 9 |
| Model | Random-effects |
| Method | DerSimonian-Laird |
| Observed | 9 |
| Imputed | 0 |
| Pooling |  |
| Parameter | Value |
| Model | Random-effects |
| Method | DerSimonian-Laird |
| Results |  |
| Studies | Effect size (95% CI) |
| Observed | 23.637% (95% CI: 15.331, 31.943%) |
| Observed + Imputed | 23.637% (95% CI: 15.331, 31.943%) |

**Abbreviations:** CI=Confidence Interval

**Table 6:** Nonparametric trim-and-fill analysis of publication bias imputing on the right.

| Nonparametric trim-and-fill analysis of publication bias |  |
| --- | --- |
| Linear estimator, imputing on the right |  |
| Iteration |  |
| Parameter | Value |
| No of studies | 12 |
| Model | Random-effects |
| Method | DerSimonian-Laird |
| Observed | 9 |
| Imputed | 3 |
| Pooling |  |
| Parameter | Value |
| Model | Random-effects |
| Method | DerSimonian-Laird |
| Results |  |
| Studies | Effect size (95% CI) |
| Observed | 23.637% (95% CI: 15.331, 31.943%) |
| Observed + Imputed | 32.226% (95% CI: 17.312, 47.139%) |

**Abbreviations:** CI=Confidence Interval

### Sensitivity analysis of ESBL-PE colonization rate

Sensitivity analysis was carried out using leave-one-out approach to detect any potential outlier studies. Based on the random effects model, no single study had a disproportionate impact on the overall pooled estimates of ESBL-PE colonization rates. The findings revealed that the estimates from all included studies were within the pooled estimate’s confidence interval, indicating the reliability of the aggregated results (**Table 7 and S3 Fig**).

**Table 7:** Sensitivity analysis of ESBL-PE colonization rate among Food handlers in Africa, 2025.

| Sensitivity analysis of ESBL-PE |  |  |  |
| --- | --- | --- | --- |
| Study omitted | Estimate | 95% Confidence Interval | P-value |
| Ahmed et al, 2023 [38] | 23.82 | 15.05, 32.59 | <0.001 |
| Amare et al, 2022 [39] | 23.92 | 14.93, 32.91 | <0.001 |
| Aworh et al, 2020 [40] | 24.60 | 15.65, 33.55 | <0.001 |
| Diriba et al, 2020 [41] | 24.56 | 15.49, 33.63 | <0.001 |
| Godonou et al, 2019 [26] | 17.23 | 10.11, 24.36 | <0.001 |
| Juma et al, 2017 [24] | 26.42 | 17.01, 35.83 | <0.001 |
| Mwanginde et al, 2021 [42] | 22.01 | 13.64, 30.37 | <0.001 |
| Sallem et al, 2012-2017 [43] | 24.36 | 14.41, 34.32 | <0.001 |
| Sanneh et al, 2015 [25] | 26.27 | 16.42, 36.13 | <0.001 |
| **Pooled** | **23.64** | **15.3, 31.94** |  |

**Abbreviations:** CI=Confidence Interval; ESBL-PE=Extended-spectrum β-lactamase-producing *Enterobacteriaceae*.

## Discussion

Extended-spectrum β-lactamase-producing *Enterobacteriaceae* (ESBL-PE) represent a critical public health crisis, driven by their rapidly increasing incidence and their efficient transmission of resistance genes to other bacterial species. This resistance renders the powerful third-generation cephalosporin antibiotics ineffective, severely limiting treatment options and leading to longer hospital stays, higher healthcare costs, and increased mortality [3].

This meta-analysis revealed that nearly one in four, 23.64% (95% CI: 15.3, 31.94%) food handlers in Africa are colonized with ESBL-PE. This finding is consistent with other high-risk groups, such as residents of long-term care facilities and patients with hematological malignancies, where colonization rates of 18% and 19% have been reported, respectively [27, 28]. It also aligns closely with a previous review from Ethiopia, which reported an overall prevalence of 28.5% across all studied groups [29]. The high prevalence identified in our study indicates a substantial community-wide dissemination of ESBL-PE across Africa. This is likely driven by high rates of antibiotic consumption, particularly beta-lactams, which promotes the selection and spread of resistance genes [30].

Notably, the pooled colonization rates of ESBL-PE in the current meta-analysis is slightly lower than rates reported in hospitalized populations in Sub-Saharan Africa (32%) [19], and globally (45.6%) [18]. This variation is might be because of using hospitalized patients as a study subjects who have frequent contact with healthcare environments and receive more antibiotics, increasing their colonization risk [31]. Conversely, our result is higher than the 14% colonization rate reported in a global meta-analysis from healthy community populations [32]. This variation might be because of that food handlers represent a sub-population with a higher risk of exposure than the general population [33]. Furthermore, regional factors in Africa, such as differing antibiotic prescription practices and sanitation infrastructure, may contribute to a higher community-level burden of ESBL-PE compared to other parts of the world [34].

This review identified a high degree of heterogeneity among the included studies (I² = 98.33%, *p* <0.001). To explore this, a subgroup analysis by region was performed, which revealed that Western Africa had the highest pooled prevalence of ESBL-PE colonization rate (33.3%). This elevated rate may be influenced by the nature of the study subjects in this subgroup; two of the three studies from this region focused on slaughterhouse workers, a subgroup of food handlers with particularly high occupational exposure. Their frequent contact with live animals and raw meat, which are known reservoirs for ESBL-PE, suggesting a heightened risk of occupational exposure [35]. Conversely, the lowest pooled colonization rate (19.28%) was found in Eastern Africa. This lower prevalence may be partially explained by the inclusion of a study conducted among hotel food service workers, who are typically less involved in the high-risk handling of raw animal products compared to other food handlers, potentially reducing their frequency of exposure [24].

According to the subgroup analysis by study period, the pooled prevalence of fecal ESBL-PE was greater between 2012 and 2019 (25.08%) than between 2020 and 2025 (22.53%). This decline in the latter period might be due to the widespread implementation of enhanced hygiene measures, including intensified handwashing, and surface disinfection during the COVID-19 (Corona Virus Disease-19) pandemic. These practices likely reduced the general transmission of pathogens, including ESBL-PE, within community and occupational settings [36].

Furthermore, a subgroup analysis based on methods of ESBL-PE detection revealed that the highest colonization rate was reported by studies that used the DDST (25.06%) than the CDT method (19.40%). The higher sensitivity of the DDST, potentially due to its ability to better detect weak ESBL producers and a broader range of ESBL types, likely contributes to the higher colonization rate observed in studies using this method [37].

## Limitations

This study has several limitations. First, the search strategy may not have captured all relevant literature, as it was restricted to three databases, which limited the number of studies for the meta-analysis. Second, regarding the included studies themselves, several focused exclusively on single bacterial species, such as ESBL-producing *E. coli* or *K. pneumoniae*; this narrow scope likely led to an underestimation of the true colonization rate. Finally, insufficient data were available on risk factors, preventing a meaningful analysis of potential contributors to ESBL-PE colonization among food handlers in Africa.

## Conclusion and recommendations

This meta-analysis, reveals that approximately one in four food handlers in Africa is colonized with ESBL-PE. This high prevalence of colonization poses a significant risk, as it can lead to difficult-to-treat infections and facilitates the silent spread of resistance within communities. The considerable heterogeneity observed among the studies suggests that this burden may vary significantly across different regions and settings. Therefore, we recommend coordinated, multi-sectoral interventions. These should include the implementation of regular screening and antimicrobial stewardship programs targeted at food handlers, alongside enhanced infection prevention and control measures in food establishments.

## Data Availability

All data produced in the present work are contained in the manuscript

## Declarations

### Ethics approval and consent to participate

Since this study was based on data extracted from previously published studies, ethical approval was not applicable

### Consent for publication

Not applicable

## Acknowledgments

We would like to acknowledge all the authors of the original studies (including its study participants) included in our systematic review and meta-analysis. We also acknowledge the College of Medicine and Health Sciences, University of Gondar

## Disclosure statement

The authors declare that they have no conflicting interests.

## Funding

No funding was provided for this study.

## Data availability statement

All relevant data are available within the manuscript or supplementary materials; for further information, please contact the corresponding author.

## Abbreviations

AMR: Antimicrobial Resistance
CDC: Centers for Disease Control and Prevention
CDT: Combination Disk Test
CI: Confidence Interval
CLSI: Clinical and Laboratory Standards Institute
CoCoPop: Condition, Context and Population
DDST: Double Disk Synergy Test
ESBL: Extended Spectrum Beta-Lactamase
ESBL-PE: Extended Spectrum Beta-Lactamase-Producing Enterobacteriaceae
JBI: Joanna Briggs Institute
MDR: Multidrug Resistance
MeSH: Medical Subject Heading
PRISMA: Preferred Reporting Items for Systematic Reviews and Meta-Analysis
SSA: Sub-Saharan Africa
US: United States
WHO: World Health Organization

## Authors’ contributions

Conceptualization: Amanuale Zayede, Mucheye Gizachew.

Data curation: Amanuale Zayede, Mucheye Gizachew.

Formal analysis: Amanuale Zayede, Sirak Biset, Eshet Gebrie.

Funding acquisition: Amanuale Zayede.

Investigation: Amanuale Zayede.

Methodology: Amanuale Zayede, Mucheye Gizachew, Sirak Biset, Yitayih Wondimeneh.

Project administration: Amanuale Zayede.

Resources: Amanuale Zayede.

Software: Amanuale Zayede, Mucheye Gizachew, Sirak Biset, Yitayih Wondimeneh, Eshet Gebrie.

Supervision: Amanuale Zayede, Mucheye Gizachew, Sirak Biset, Yitayih Wondimeneh, Eshet Gebrie, Mitkie Tigabie, Henok Worku, Biruktawit Abebe, Haymanot Til.

Validation: Amanuale Zayede, Mucheye Gizachew, Sirak Biset, Yitayih Wondimeneh, Eshet Gebrie, Mitkie Tigabie, Henok Worku, Biruktawit Abebe, Haymanot Til.

Visualization: Amanuale Zayede, Mucheye Gizachew, Sirak Biset, Yitayih Wondimeneh, Eshet Gebrie, Mitkie Tigabie, Henok Worku, Biruktawit Abebe, Haymanot Til.

Writing- original draft: Amanuale Zayede.

Writing-review & editing: Amanuale Zayede, Mucheye Gizachew, Sirak Biset, Yitayih Wondimeneh.

## Supporting information

**S1 Table:** PRISMA checklist, 2020

**S2 Table:** PubMed Search strategy

**S1 Fig:** Forest plot of ESBL-PE other than *E.coli* and *Klebsiella* species

**S2 Figs:** Forest plot of subgroup analysis

**S3 Fig:** Forest plot of sensitivity analysis

**S4 Figs:** Funnel plot of trim and fill analysis

## Notes

### Competing Interest Statement

The authors have declared no competing interest.

### Clinical Protocols

https://www.crd.york.ac.uk/PROSPERO/

### Funding Statement

This study did not receive any funding

## References

[1] J. D. Pitout and K. B. Laupland, "Extended-spectrum β-lactamase-producing Enterobacteriaceae: an emerging public-health concern," The Lancet infectious diseases, vol. 8, no. 3, pp. 159–166, 2008.

[2] C. Scully, J. L. Posse, and P. D. Dios, Saliva protection and transmissible diseases. Academic Press, 2017.

[3] World Health Organization, WHO bacterial priority pathogens list, 2024: bacterial pathogens of public health importance, to guide research, development, and strategies to prevent and control antimicrobial resistance. World Health Organization, 2024.

[4] D. L. Paterson, "Resistance in gram-negative bacteria: Enterobacteriaceae," American Journal of Infection control, vol. 34, no. 5, pp. S20–S28, 2006.

[5] G. H. Talbot, J. Bradley, J. E. Edwards Jr, D. Gilbert, M. Scheld, and J. G. Bartlett, "Bad bugs need drugs: an update on the development pipeline from the Antimicrobial Availability Task Force of the Infectious Diseases Society of America," Clinical Infectious Diseases, vol. 42, no. 5, pp. 657–668, 2006.

[6] S. Ajulo and B. Awosile, "Global antimicrobial resistance and use surveillance system (GLASS 2022): Investigating the relationship between antimicrobial resistance and antimicrobial consumption data across the participating countries," PLoS One, vol. 19, no. 2, p. e0297921, 2024.

[7] S. Kariuki, "Global burden of antimicrobial resistance and forecasts to 2050," The Lancet, vol. 404, no. 10459, pp. 1172–1173, 2024.

[8] A. Godonou et al., "High faecal carriage of extended-spectrum beta-lactamase producing Enterobacteriaceae (ESBL-PE) among hospitalized patients at Sylvanus Olympio Teaching Hospital, Lomé, Togo in 2019," African Journal of Clinical and Experimental Microbiology, vol. 23, no. 1, pp. 40–48, 2022.

[9] P. A. Bradford, "Extended-spectrum β-lactamases in the 21st century: characterization, epidemiology, and detection of this important resistance threat," Clinical Microbiology Reviews, vol. 14, no. 4, pp. 933–951, 2001.

[10] M. Asma and A. Jasser, "Extended-spectrum beta-lactamases [ESBLs]: a Global Problem," 2006.

[11] M. C. Basavaraj, P. Jyothi, and P. V. Basavaraj, "The prevalence of ESBL among Enterobacteriaceae in a tertiary care hospital of North Karnataka, India," Journal of Clinical and Diagnostic Research, vol. 5, no. 3, pp. 470–475, 2011.

[12] C. f. D. Control and Prevention, Antibiotic resistance threats in the United States, 2019. *US Department of Health and Human Services, Centres for Disease Control* and …, 2019.

[13] W. Elliott and J. Chan, "Plazomicin injection (Zemdri)," Internal Medicine Alert, vol. 40, no. 15, 2018.

[14] G. L. Daikos et al., "Prospective observational study of the impact of VIM-1 metallo-β-lactamase on the outcome of patients with Klebsiella pneumoniae bloodstream infections," Antimicrobial Agents and Chemotherapy, vol. 53, no. 5, pp. 1868-1873, 2009.

[15] A. J. Stewardson et al., "The health and economic burden of bloodstream infections caused by antimicrobial-susceptible and non-susceptible Enterobacteriaceae and Staphylococcus aureus in European hospitals, 2010 and 2011: a multicentre retrospective cohort study," Eurosurveillance, vol. 21, no. 33, p. 30319, 2016.

[16] K. G. Jarvis et al., "Microbiomes associated with foods from plant and animal sources," Frontiers In Microbiology, vol. 9, p. 2540, 2018.

[17] S. Vergara-López, M. Domínguez, M. Conejo, Á. Pascual, and J. Rodríguez-Baño, "Wastewater drainage system as an occult reservoir in a protracted clonal outbreak due to metallo-β-lactamase-producing Klebsiella oxytoca," Clinical Microbiology and Infection, vol. 19, no. 11, pp. E490-E498, 2013.

[18] D. Abera, A. Alemu, A. Mihret, A. A. Negash, W. E. Abegaz, and K. Cadwell, "Colonization with extended spectrum beta-lactamase and carbapenemases producing Enterobacteriaceae among hospitalized patients at the global level: A systematic review and meta-analysis," PLoS One, vol. 18, no. 11, p. e0293528, 2023.

[19] J. M. Lewis, R. Lester, P. Garner, and N. A. Feasey, "Gut mucosal colonisation with extended-spectrum beta-lactamase producing Enterobacteriaceae in sub-Saharan Africa: a systematic review and meta-analysis," Wellcome Open Research, vol. 4, p. 160, 2020.

[20] N. R. Haddaway, M. J. Page, C. C. Pritchard, and L. A. McGuinness, "PRISMA 2020: An R package and Shiny app for producing PRISMA 2020-compliant flow diagrams, with interactivity for optimised digital transparency and Open Synthesis," Campbell Systematic Reviews, vol. 18, no. 2, p. e1230, 2022.

[21] Z. Munn, S. Moola, K. Lisy, D. Riitano, and C. Tufanaru, "Methodological guidance for systematic reviews of observational epidemiological studies reporting prevalence and cumulative incidence data," JBI Evidence Implementation, vol. 13, no. 3, pp. 147–153, 2015.

[22] M. Borenstein, H. Cooper, L. Hedges, and J. Valentine, "Heterogeneity in meta-analysis," The Handbook of Research Synthesis and Meta-analysis, vol. 3, pp. 453–70, 2019.

[23] M. J. Page, J. A. Sterne, J. P. Higgins, and M. Egger, "Investigating and dealing with publication bias and other reporting biases in meta-analyses of health research: A review," Research Synthesis Methods, vol. 12, no. 2, pp. 248–259, 2021.

[24] A. O. Juma, "Antimicrobial susceptibility profiles and genotypic characterization of selected Enterobacteriaceae strains isolated from food handlers in Nairobi, Kenya," COHES-JKUAT, 2017.

[25] B. Sanneh et al., "Prevalence and risk factors for faecal carriage of Extended Spectrum β-lactamase producing Enterobacteriaceae among food handlers in lower basic schools in West Coast Region of The Gambia," PLoS One, vol. 13, no. 8, p. e0200894, 2018.

[26] A. M. Godonou et al., "High carrying rate of extended-spectrum beta-lactamase (ESBL) producing Enterobacteriaceae by slaughterhouse workers in Lomé, Togo in 2019," Microbiol Res J Int, pp. 30–41, 2020.

[27] M. Alevizakos, S. Karanika, M. Detsis, and E. Mylonakis, "Colonisation with extended-spectrum β-lactamase-producing Enterobacteriaceae and risk for infection among patients with solid or haematological malignancy: a systematic review and meta-analysis," International Journal of Antimicrobial Agents, vol. 48, no. 6, pp. 647–654, 2016.

[28] M. E. Flokas, M. Alevizakos, F. Shehadeh, N. Andreatos, and E. Mylonakis, "Extended-spectrum β-lactamase-producing Enterobacteriaceae colonisation in long-term care facilities: a systematic review and meta-analysis," International Journal of Antimicrobial Agents, vol. 50, no. 5, pp. 649–656, 2017.

[29] M. Tigabie, et al., "Colonization with extended-spectrum β-lactamase and carbapenemase-producing Enterobacterales in Ethiopia: A systematic review and meta-analysis," PLoS One, vol. 20, no. 4, p. e0316492, 2025.

[30] O. G. Onduru, R. S. Mkakosya, S. Aboud, and S. F. Rumisha, "Genetic determinants of resistance among ESBL-producing enterobacteriaceae in community and hospital settings in east, central, and Southern Africa: A systematic review and meta-analysis of prevalence," Canadian Journal of Infectious Diseases and Medical Microbiology, vol. 2021, no. 1, p. 5153237, 2021.

[31] J. Bizimana, J. Ndayisenga, H. Kajumbura, P. Mulepo, and N. F. Christine, "Colonization of patients hospitalized at orthopedic department of tertiary hospital in Uganda with extended-spectrum beta-lactamase-producing enterobacterales," (in eng), Antimicrob Resist Infect Control, vol. 12, no. 1, p. 26, Apr 1 2023.

[32] S. Karanika, T. Karantanos, M. Arvanitis, C. Grigoras, and E. Mylonakis, "Fecal colonization with extended-spectrum beta-lactamase–producing Enterobacteriaceae and risk factors among healthy individuals: a systematic review and metaanalysis," Reviews of Infectious Diseases, vol. 63, no. 3, pp. 310–318, 2016.

[33] A. J. Stewardson et al., "Extended-Spectrum β-Lactamase–Producing Enterobacteriaceae in Hospital Food: A Risk Assessment," Infection Control & Hospital Epidemiology, vol. 35, no. 4, pp. 375–383, 2014.

[34] M. Garé et al., "Antimicrobial resistance of Enterobacterales in Central Africa: a systematic review and meta-analysis," Communications Medicine, vol. 5, no. 1, p. 453, 2025/11/04 2025.

[35] C. Agyare, V. E. Boamah, C. N. Zumbi, and F. B. Osei, "Antibiotic use in poultry production and its effects on bacterial resistance," in Antimicrobial resistance-A global threat: IntechOpen, 2018.

[36] D. L. Monnet and S. Harbarth, "Will coronavirus disease (COVID-19) have an impact on antimicrobial resistance?," Eurosurveillance, vol. 25, no. 45, p. 2001886, 2020.

[37] P. Das, D. Mahapatra, and S. Mazumder, "A Guide Towards the Phenotypic Detection of Extended-spectrum β-lactamases Production in Enterobacteriaceae: Alone or in Presence of Other Interfering Enzymes," J Pure Appl Microbiol, vol. 17, no. 3, pp. 1410–21, 2023.

[38] H. A. Ahmed et al., "Extended-spectrum β-lactamase-producing E. coli from retail meat and workers: genetic diversity, virulotyping, pathotyping and the antimicrobial effect of silver nanoparticles," BMC Microbiology, vol. 23, no. 1, p. 212, 2023.

[39] A. Amare, S. Eshetie, D. Kasew, and F. Moges, "High prevalence of fecal carriage of Extended-spectrum beta-lactamase and carbapenemase-producing Enterobacteriaceae among food handlers at the University of Gondar, Northwest Ethiopia," PloS One, vol. 17, no. 3, p. e0264818, 2022.

[40] M. K. Aworh, O. Abiodun-Adewusi, N. Mba, B. Helwigh, and R. S. Hendriksen, "Prevalence and risk factors for faecal carriage of multidrug resistant Escherichia coli among slaughterhouse workers," Scientific Reports, vol. 11, no. 1, p. 13362, 2021.

[41] K. Diriba, E. Awulachew, L. Tekele, and Z. Ashuro, "Fecal carriage rate of extended-spectrum beta-lactamase-producing Escherichia coli and Klebsiella pneumoniae among apparently health food handlers in Dilla University student cafeteria," Infection and Drug Resistance, pp. 3791-3800, 2020.

[42] L. W. Mwanginde, M. Majigo, D. C. Kajeguka, and A. Joachim, "High Carriage Rate of Extended-Spectrum β-Lactamase-Producing Escherichia coli and Klebsiella Species among Poultry Meat Vendors in Dar es Salaam: The Urgent Need for Intervention to Prevent the Spread of Multidrug-Resistant Pathogens," International Journal of Microbiology, vol. 2021, no. 1, p. 6653993, 2021.

[43] N. Sallem, A. Hammami, and B. Mnif, "Trends in human intestinal carriage of ESBL-and carbapenemase-producing Enterobacterales among food handlers in Tunisia: emergence of C1-M27-ST131 subclades, bla OXA-48 and bla NDM," Journal of Antimicrobial Chemotherapy, vol. 77, no. 8, pp. 2142–2152, 2022.

